# The testing of the DAC algorithm for classifying Lymphoma Cell of Origin samples using HTG’s targeted gene expression system

**DOI:** 10.1101/2022.04.14.22270303

**Authors:** John R. Davies, Matthew A. Care, Tracey Mell, Sharon Barrans, Cathy Burton, David R. Westhead

**Author notes:** **Competing interests** None.

## Abstract

An implementation of the windows based DLBCL automatic classification (DAC) algorithm for determining cell of origin (COO) written in the R language and designed for use with the HTG EdgeSeq Reveal Software package is described. Classifications using the new implementation (DAC-R) were compared to the those using the original version (DAC-Win), the HTG DLBCL COO classifier, the HTG EdgeSeq Reveal DLBCL algorithm. Classifications made using HTG data were tested for concordance with those made using WG-DASL data for the same samples. To analyse small numbers of samples a background dataset was developed to allow for the reliable classification of individual cases. Classification accuracy was assessed using percentage of agreement of discrete classification groups and Pearson correlation of probability scores where appropriate. In the data tested it was seen that the correlation of probability scores for DAC-R and DAC-Win was 0.9985 or higher. Agreement with the HTG DLBCL COO classifier was 85.1% and agreement with the HTG EdgeSeq Reveal DLBCL algorithm was also high (Pearson correlation of GCB probabilities = 0.91). Agreement between classifications made using HTG and WG-DASL data was also high (83.7%). In summary, the R based algorithm of DAC successfully replicates the functionality of the original routine and produces comparable COO calls in data produced from the HTG Pan B-Cell Lymphoma Panel to the other methods listed.

## Introduction

Diffuse large B-cell lymphoma (DLBCL) is the most common type of non-Hodgkin lymphoma but is heterogeneous with respect to clinical presentation and response to treatment. Some of this heterogeneity can be explained by the putative cell of origin (COO) of the tumours. COO defined by gene expression profiling is a well-established method to differentiate between lymphomas of Germinal Center B-cell (GCB) and those of Activated B-cell (ABC) origin (Alizadeh *et al*, 2000) and these two main sub-types of DLBCL have differential responses to standard treatment. GCB generally has a more favourable outcome compared to ABC following standard R-CHOP [rituximab, cyclophosphamide, doxorubicin hydrochloride, vincristine (Oncovin, Vincasar PFS), prednisolone] chemotherapy, which has been replicated in numerous studies (Lenz *et al*, 2008; Lenz *et al*, 2010; Painter *et al*, 2019). In addition, there is a third DLBCL type known as unclassified (UNC) or Type III, which are generally associated with high levels of necrosis, stroma or infiltrating T cells (Alizadeh *et al*, 2000; Lenz *et al*, 2008; Care *et al*, 2015).

Gene expression profiling can now be applied to routinely processed formalin-fixed paraffin-embedded (FFPE) diagnostic tissue biopsies but, despite this, it has not been widely incorporated into routine clinical use, and the surrogate immunohistochemistry (IHC)-based Hans test remains as standard practice. The Hans test uses just three markers (CD10, BCL6 and IRF4/MUM1) to classify patient samples as either GCB or non-GCB; however, reproducibility has proved difficult, and this classification does not identify significant differences in overall survival (Read et al 2014). COO classification was recognised in the 2017 update of the World Health Organization classification of lymphoid neoplasms (Swerdlow *et al*, 2016), which states that COO should be defined preferably by GEP, but recommends Hans IHC only where this is not possible.

The HTG assay is a targeted gene expression test using a quantitative Nuclease Protection Assay (qNPA) which enables low quantities of formalin-fixed paraffin embedded (FFPE) material to be assayed with a streamlined workflow. There are two panels available which contain probes targeting genes specifically of interest in lymphoma. One such qNPA panel is the HTG DLBCL COO panel which is designed to provide a COO classification for DLBCL. It is CE-IVD marked for use as an *in vitro* diagnostic for the classification of samples as GCB or ABC, with cases that do not fall into either category categorized as UNC (Xu-Monette *et al, 2015*). The software associated with this HTG DLBCL COO panel incorporates a DLBCL COO classifier which provides a classification and probability score for GCB, ABC and UNC. A second qNPA panel is the HTG Pan B-Cell Lymphoma Panel which can assay a much broader set of genes than the DLBCL COO panel. It is able to assay 298 mRNAs which are considered important in lymphoma. Within the HTG EdgeSeq Reveal software, there is an algorithm which can use PanB gene expression profiles to give a probability score that a sample is GCB (Xu-Monette *et al*, 2015; Schaffer *et al*, 2018). However, the HTG EdgeSeq Reveal DLBCL algorithm does not currently provide the full COO classification.

The freely available DLBCL automatic classification (DAC) algorithm is a sophisticated, platform agnostic algorithm which is able to effectively classify DLBCL samples into GCB, ABC and UNC groups using gene expression data and provides a probability score for each class (Care *et al*, 2013). DAC is a meta-classifier using a combination of 4 different machine learning tools. A rigorous cross comparison examining the COO classification of DLBCL showed a high level of concordance between COO classifications provided by DAC produced using four different gene expression platforms (Affymetrix and Illumina DASL microarray gene expression, RNAseq) as well as the HTG DLBCL COO panel and classifier (Ahmed *et al, 2021*).

The original DAC classifier (DAC-Windows) has been coded as an R version of the DAC classifier in order to facilitate integration of the algorithm into external software, specifically the HTG EdgeSeq Reveal Software package. The recoded R implementation of DAC is known as DAC-R.

The purpose of the paper is to describe the testing of this DAC-R implementation and extend the testing of DAC by assessing the algorithm’s performance in a series of comparisons using multiple datasets.

## Materials and Methods

The R implementation of DAC has been written in R version 3.6.2 using the RWeka package to replicate the original classifier as described in Care *et al*. (2013). Briefly, the classifier is a composite of four algorithms within Weka (LMT_J48_RF100_SMO) trained upon a dataset of 240 DLBCL samples generated by Wright *et al*. (2003) across 20 genes. Classification is performed for all four methods and a final result is generated by averaging the probabilities. The classification that has the highest resultant score is the final classification call.

The samples used were FFPE samples obtained from the REMoDL-B trial (Davies *et al*, 2019), the HMRN population-based series (Painter *et al*, 2019) and the MaPLe Study. The MaPLe study is an ongoing multicenter prospective observational cohort study conducted by the Southamption clinical trials unit concerning patients with suspected or confirmed diffuse large B cell lymphoma (University of Southampton, 2021). Table 2 shows how each study contributes to each test.

**Table 1:** A summary of the algorithms and datasets used in this study. Materials and Methods

| Algorithm | Dataset compatibility | Output | Regulatory status |
| --- | --- | --- | --- |
| **DAC - Windows** | Microarray (including DASL)<br>RNA Seq<br>HTG Pan B-Cell<br>Lymphoma Panel | Probability of GCB<br>Probability of ABC<br>Probability of UNC<br>Classification | Research Use Only |
| **DAC-R** | Microarray<br>RNA Seq<br>HTG Pan B-Cell<br>Lymphoma Panel | Probability of GCB<br>Probability of ABC<br>Probability of UNC<br>Classification | Research Use Only |
| **HTG DLBCL COO classifier** | HTG DLBCL COO panel | Probability of GCB<br>Probability of ABC<br>Probability of UNC<br>Classification | IVD CE Marked |
| **HTG EdgeSeq Reveal DLBCL algorithm** | HTG Pan B-Cell<br>Lymphoma Panel | Probability of GCB | Research Use Only |

**Table 2.** Summary of samples used for evaluating performance of the DAC-R algorithm.

|  | DAC-R v DAC - Windows | Comparison of DAC-R v HTG DLBCL COO | DAC-R v HTG EdgeSeq Reveal | DAC-R on HTG PanB v WG-DASL (DAC) | Background validation |
| --- | --- | --- | --- | --- | --- |
| **REMoDL-B** | 86 | 54 | 38 | 86 | - |
| **HMRN population** | 100 | - | 29 | - | - |
| **MaPLE** | 57 | - | - | - | 57 |
| **Total** | 243 | 54 | 67 | 86 | 57 |

The HTG EdgeSeq assays were run using the protocols as provided by the manufacturer (HTG Molecular Diagnostics, Tucson, USA). Briefly, FFPE samples were heated with Lysis buffer for 95 ^°^C for 15 minutes, Proteinase K was added and incubated at 50 ^°^C for 3 hours. Following the qNPA in the EdgeSeq processor, the resulting probe DNA was amplified using PCR, cleanup carried out using AMPure beads and the library quantities determined using quantitative PCR. The libraries were pooled in equivalent quantities and run on an Illumina MiSeq instrument following manufacturer’s recommendation (Illumina, San Diego, USA). Quantitation of the reads from the Fastq files used the HTG EdgeSeq parser software. Samples that did not pass two out of four quality control checks (negative/positive ratio, maximum raw read count, median read count and coefficient of variation) were excluded. Principal component analysis was also conducted on the transformed data as a further test of sample validity within the cohort, and extreme outliers manually removed from the data prior to classification. Log transformation of count data was conducted using the “rlog” function of DESeq2 from the DESeq2 package in R version 3.6.2.

DAC is a classifier that has been designed to use data generated from different gene expression technologies. Consequently, it requires a background data set from the same platform to normalize the data of interest. If a large batch of samples is to be classified the data set can act as its own background for normalization purposes (batch mode). Unless otherwise stated, the batch mode was used in this study. Alternatively, individual samples can be classified using a previously defined background data set from the same platform for normalization (standalone mode). We describe the selection and testing of different background datasets and compare performance based on comparison of COO calls and related probability scores with previously classified datasets.

Tables and scatterplots were used to compare classification results of DAC-R-with DAC -Windows, HTG DLBCL COO classifier, HTG EdgeSeq Reveal DLBCL algorithm GCB score and DAC-windows using whole genome DASL data (WG-DASL). LOESS curves were fitted to show the relationship between probability scores. Correlation of probability scores was tested using Pearson’s correlation coefficient.

## Results

### Comparison of DAC-R in Batch mode versus DAC Windows implementation

The first step was to compare the classification obtained using the new R implementation of DAC (DAC-R) with the classification obtained using the original Windows-based DAC implementation. This comparison was carried out using 243 samples generated with the HTG Pan B-Cell Lymphoma Panel. As shown in Table 3, the classifications were very similar. Out of the 243 samples, 239 were classified the same (98.4%). Two samples classified as ABC or GCB with the original Windows implementation were classified as UNC with DAC-R. One ABC sample with the Windows version was called as GCB with DAC-R and one GCB with windows was called ABC with DAC-R. These changes of classification were related to small variations in probabilities between the two versions of classifier (0.009-0.033) in cases where the probability values for ABC and GCB were very similar (edge cases), resulting in a shift of the call. This has been observed previously in a comparative analysis of cell of origin classification across multiple gene expression platforms (Ahmed *et al, 2021*).

**Table 3:** Comparison of the classifications generated with the R implementation of DAC with the DAC Windows version (HTG-Win).

|  |  | DAC (HTG-R) |  |  |  |
| --- | --- | --- | --- | --- | --- |
|  |  | ABC | GCB | UNC | Total |
| DAC<br>(HTG-Win) | ABC | 66 | 1 | 1 | 68 |
|  | GCB | 1 | 151 | 1 | 153 |
|  | UNC | 0 | 0 | 22 | 22 |
|  | Total | 67 | 152 | 24 | 243 |

As well as comparing the classifications, we also sought to compare the probability scores of each of the three DLBCL subtypes. The Pearson correlation coefficient of the GCB probability score generated with the Windows DAC compared to the GCB probability produced by the DAC-R very high at 0.995. With the ABC probability scores the correlation was 0.995 and the probability score of UNC between the two implementations was 0.9985 (Figure 1). These results confirm that the implementation of the DAC in R can generate near identical results to the original Windows based implementation.

**Figure 1.**
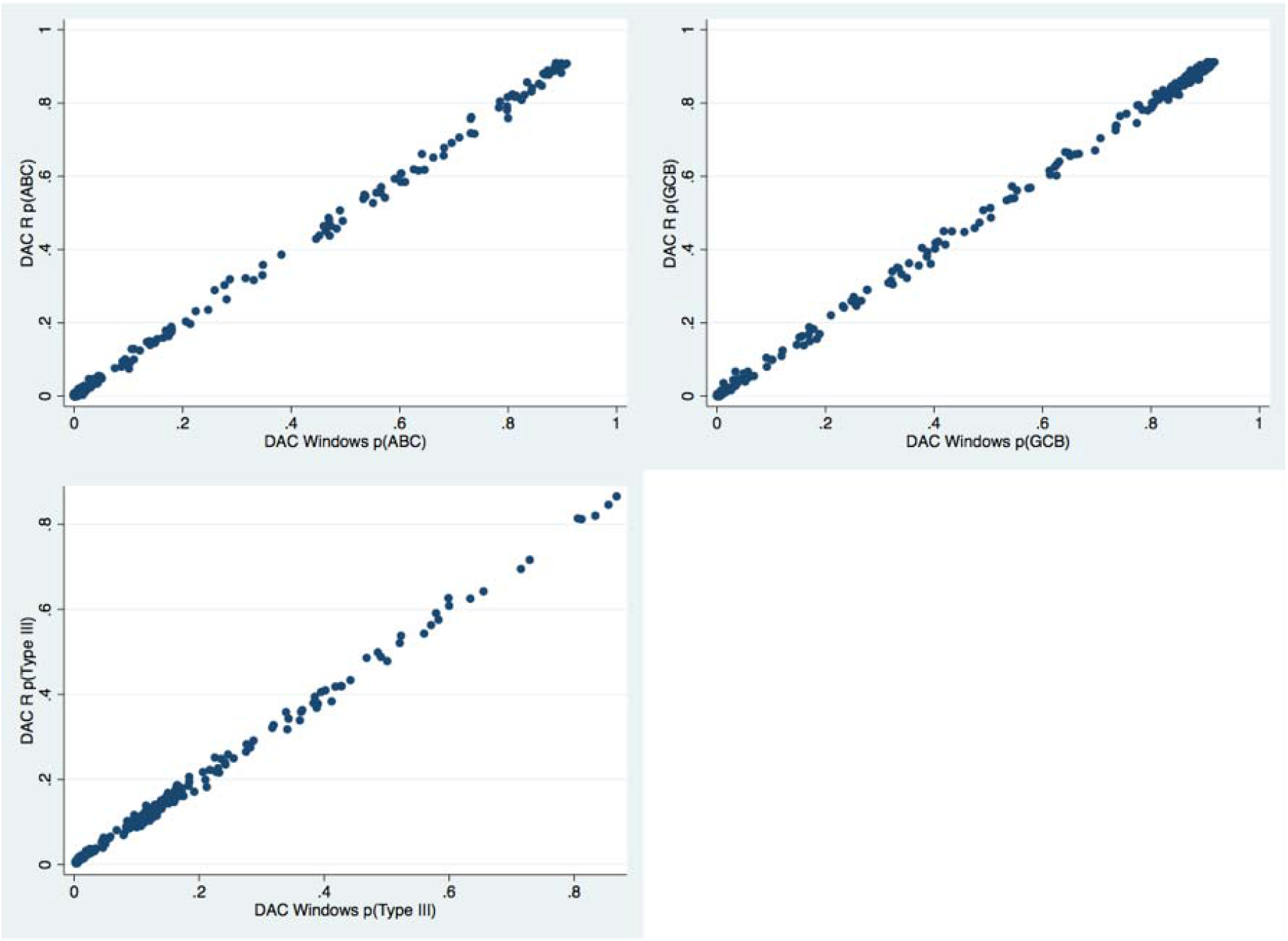
Comparison of the probability scores generated by the two DAC implementations. Comparison of DAC-R in Batch mode with the HTG DLBCL COO classifier.

With data generated from 54 samples using the HTG DLBCL COO panel, we compared COO classifications using DAC-R with classifications generated using the HTG DLBCL COO classifier (Ahmed *et al*, 2021). From the confusion table (Table 4), 46 of the 54 samples (85.1%) were classified the same. Of the 8 discrepant classifications, 5 samples were classified as UNC with the DAC-R. However, the HTG DLBCL COO classifier has been designed to minimise UNC and this design likely reflects this. Of the remaining 3 discrepant samples, 2 showed borderline classification probabilities.

**Table 4:**
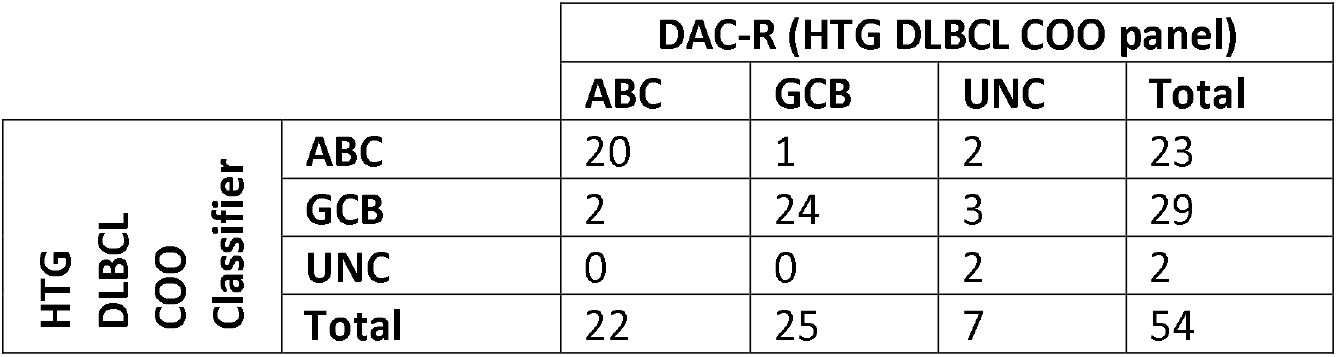
Comparison of the classification with DAC-R) and the HTG DLBCL COO classifier using gene expression profiles generated with the HTG DLBCL COO panel.

### Comparison of HTG DAC-R in Batch mode with the HTG EdgeSeq Reveal DLBCL algorithm (pGCB)

Using 67 gene expression profiles produced with the HTG Pan B-Cell Lymphoma Panel, the probability GCB score generated by DAC-R was compared to the probability GCB score from the HTG EdgeSeq REVEAL DLBCL algorithm. The data is presented in Figure 2. It was not expected that complete concordance between the two scores would be achieved since, unlike p(GCB) in HTG EdgeSeq REVEAL DLBCL, p(GCB) in DAC is also dependent on the strength of signal for p(ABC) and p(UNC). Nevertheless, there was good concordance between the two GCB probability scores (Pearson correlation =0.91), suggesting that these measures are performing similarly.

**Figure 2.**
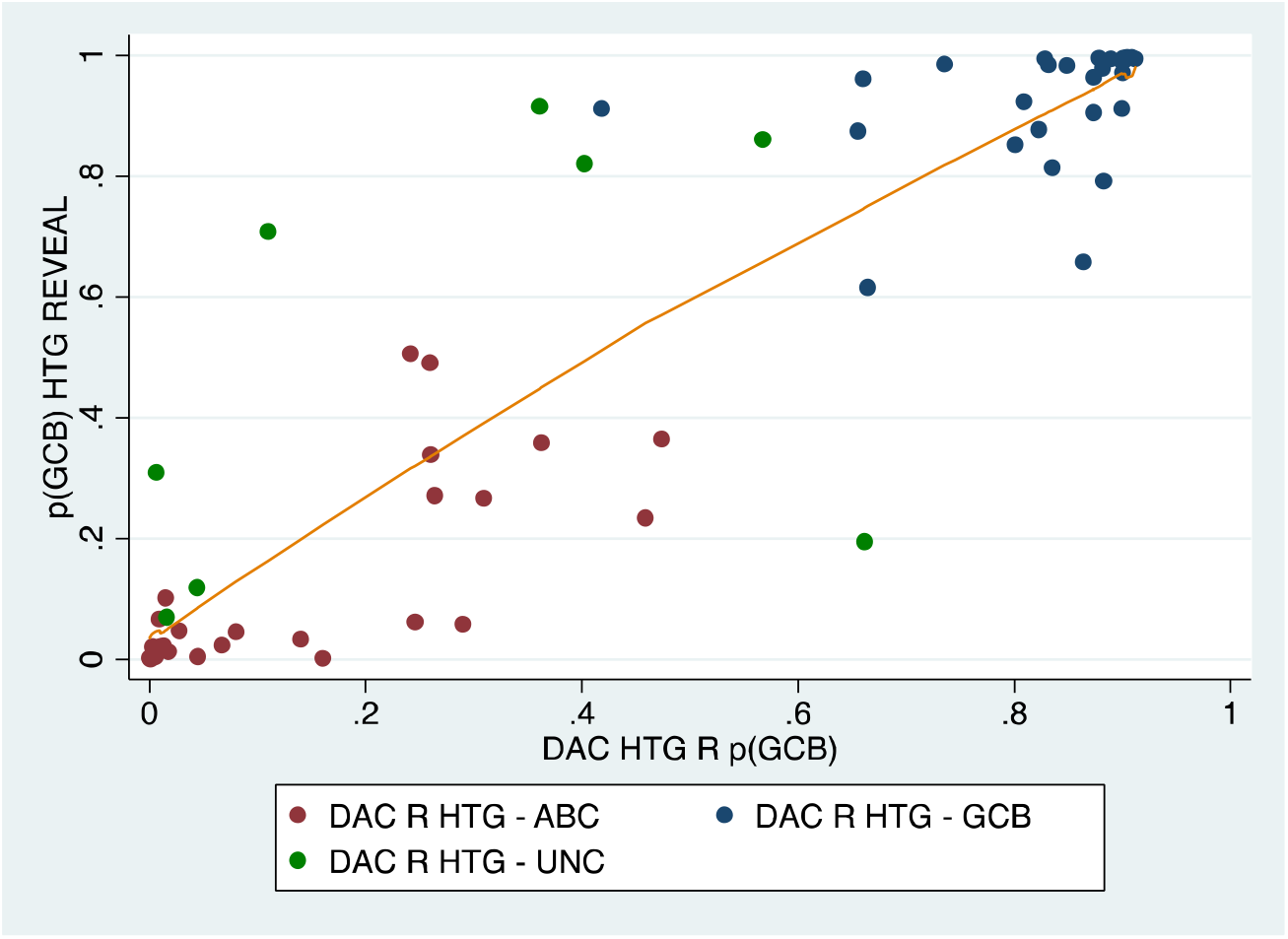
Graph of the GCB probability scores obtained using the DAC-R and the HTG EdgeSeq Reveal DLBCL algorithm. The colours indicate the classification from the DAC classifier.

The classifications of the samples were then compared. The HTG EdgeSeq Reveal DLBCL algorithm for the HTG Pan B-Cell Lymphoma Panel analysis currently only generates a GCB probability score. If we generate a hard classification from the HTG EdgeSeq Reveal DLBCL probability score using the most probable outcome, i.e. assuming a probability of greater than 0.5 is GCB and less than 0.5 is not GCB, there was very good concordance with the DAC-R classification. HTG EdgeSeq Reveal DLBCL and DAC-R agreed that 29 samples were not GCB and 31 were GCB, disagreeing on 7 samples. The samples which had discordant classifications between the two algorithms are shown in table 5. As seen in previous comparisons, the discrepancies were largely explained by samples classified as UNC using DAC or a borderline difference in the probabilities between GCB and ABC.

**Table 5:** Probability scores for the samples which were called differently between the HTG EdgeSeq Reveal DLBCL algorithm and the DAC-R classifier

| HT HTG EdgeSeq Reveal DLBCL | HTG-R Batch DAC |  |  |  |
| --- | --- | --- | --- | --- |
| P(GCB) | Group call (DAC-R) | P(GCB) | P(ABC) | P(UNC) |
| HTG EdgeSeq Reveal DLBCL $p(\text{GCB}) < 0.5$ | | | | |
| 0.194 | GCB | 0.661 | 0.080 | 0.259 |
| 0.364 | GCB | 0.474 | 0.464 | 0.062 |
| HTG EdgeSeq Reveal DLBCL $p(\text{GCB}) > 0.5$ | | | | |
| 0.506 | ABC | 0.241 | 0.656 | 0.102 |
| 0.911 | ABC | 0.418 | 0.429 | 0.153 |
| 0.915 | Type III / UNC | 0.361 | 0.013 | 0.627 |
| 0.708 | Type III / UNC | 0.110 | 0.764 | 0.814 |
| 0.820 | Type III / UNC | 0.402 | 0.022 | 0.575 |

### Comparison of HTG Pan B-Cell Lymphoma Panel data, analysed with DAC-R in Batch mode with Illumina WG-DASL data analysed with DAC-Windows

The next comparison was to investigate a subset of 86 cases from the REMoDL-B study (Davies *et al*, 2019). These have been interrogated with the HTG Pan B-Cell Lymphoma Panel assay and analysed with DAC-R. This data was compared with REMoDL-B gene expression profiles that was originally generated prospectively at trial enrollment using the Illumina microarray WG-DASL assay, and classified using DAC - Windows in real time, using a pre-defined background dataset. Data from the whole trial was subsequently re-analysed retrospectively using the original windows implementation of DAC in batch mode to take advantage of the larger normalization set, and these classifications were used for comparison.

The comparison between the two sets of the GCB, ABC and UNC probability scores is shown in Figure 3. The Pearson correlations for the sets of probability scores in the HTG Pan B-Cell Lymphoma Panel / DAC-R and WG-DASL / DAC-Windows were high, 0.9087, 0.8273 and 0.5488 for each of these three groups respectively, suggesting good agreement between the probabilities of DAC estimates between the two expression platforms. The classifications are shown in Table 6. Out of the 86 samples, 72 (83.7%) were in agreement. This is similar to the concordance observed between different platforms in previous studies (Ahmed *et al, 2021*). The HTG Pan B-Cell Lymphoma Panel / DAC-R was slightly more likely to call cases as ABC compared to Illumina WG-DASL / DAC-Windows. Variation in the composition of background cases as well as different platform chemistry and probe design is likely to explain this variation.

**Figure 3.**
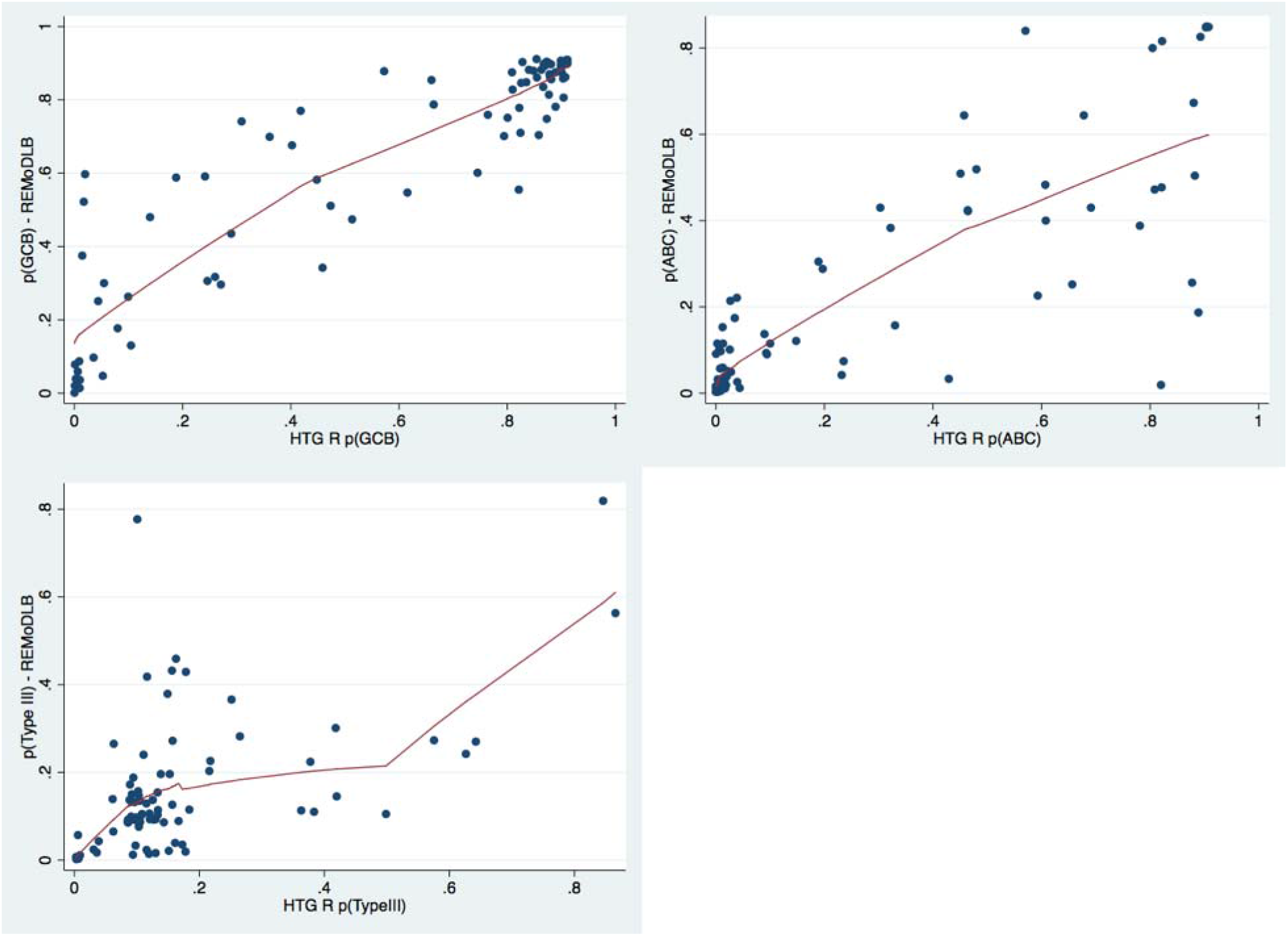
Comparing the GCB, ABC and UNC probability scores.

**Table 6:**
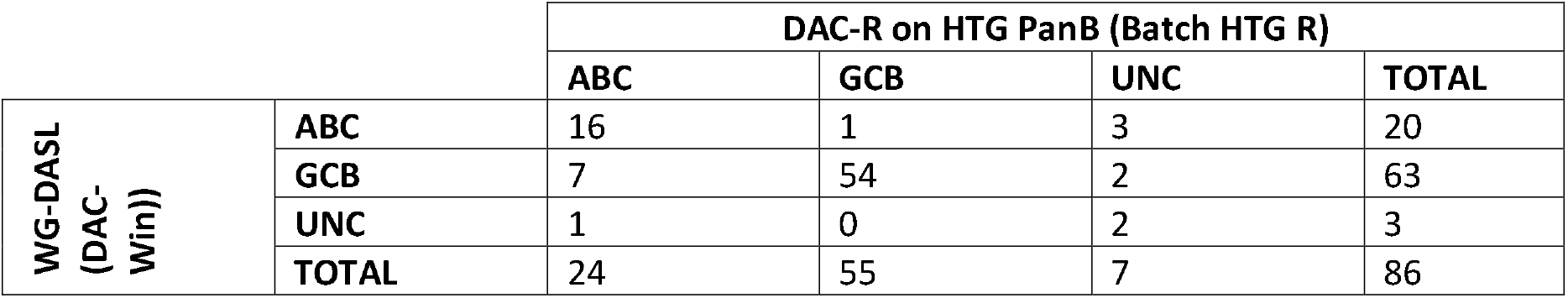
Comparison of classifications produced by the HTG Pan B-Cell Lymphoma Panel with the DAC-R compared to the WG-DASL assay using DAC-Windows REMoDL-B samples were used.

The 14 cases where the WG-DASL / DAC-Windows classify differently from the HTG Pan B-Cell Lymphoma Panel / DAC-R are shown in Table 7.

**Table 7.** Samples from the REMoDL-B trial where the call made using data generated using the HTG Pan B-Cell Lymphoma Panel (HTG-PanB) assay disagrees with the retrospective call made using data generated with the original Illumina WG-DASL data.

|  | DAC-Windows (Illumina WG-DASL) |  |  | DAC-R (HTG PanB) |  |  |  |
| --- | --- | --- | --- | --- | --- | --- | --- |
| Call | P(ABC) | P(GCB) | P(UNC) | Call | P(ABC) | P(GCB) | P(UNC) |
| ABC | 0.43 | 0.3 | 0.27 | UNC | 0.303 | 0.055 | 0.642 |
| ABC | 0.509 | 0.474 | 0.017 | GCB | 0.451 | 0.513 | 0.036 |
| ABC | 0.644 | 0.251 | 0.105 | UNC | 0.457 | 0.044 | 0.499 |
| ABC | 0.383 | 0.317 | 0.301 | UNC | 0.322 | 0.260 | 0.418 |
| GCB | 0.019 | 0.522 | 0.459 | ABC | 0.820 | 0.017 | 0.163 |
| GCB | 0.252 | 0.591 | 0.157 | ABC | 0.656 | 0.241 | 0.102 |
| GCB | 0.059 | 0.699 | 0.242 | UNC | 0.013 | 0.361 | 0.627 |
| GCB | 0.388 | 0.588 | 0.024 | ABC | 0.781 | 0.188 | 0.031 |
| GCB | 0.256 | 0.597 | 0.147 | ABC | 0.877 | 0.020 | 0.103 |
| GCB | 0.033 | 0.77 | 0.196 | ABC | 0.429 | 0.418 | 0.153 |
| GCB | 0.226 | 0.741 | 0.033 | ABC | 0.593 | 0.309 | 0.098 |
| GCB | 0.051 | 0.676 | 0.273 | UNC | 0.022 | 0.402 | 0.575 |
| GCB | 0.477 | 0.48 | 0.043 | ABC | 0.821 | 0.140 | 0.039 |
| UNC | 0.187 | 0.036 | 0.777 | ABC | 0.889 | 0.010 | 0.101 |

### Use of DAC-R in Standalone mode – Validation of a background dataset

Z-score normalisation requires estimates of the mean and standard deviation for each gene. In order to analyse small numbers of samples a separate “background” dataset is required to allow calculation of these estimates. Variation in the composition of the background dataset can have an effect on the classification of a small number of predominantly borderline cases. Specifically, if the background dataset is incorrectly balanced then, in a random sample of cases, over-represented groups will be called less frequently than expected. Intuitively the composition of the dataset should be similar to the expected proportions of the classes in the test dataset, however the composition of cases can differ between clinical trial and population-based cohorts. A comparison was carried out using 57 samples generated with the HTG Pan B-Cell Lymphoma Panel assay taken from the DLBCL MaPLe study (MaPLe | Southampton Clinical Trials Unit | University of Southampton). All samples were analysed in standalone mode one by one; first using a background derived from 34 samples generated with the HTG PanB qNPA assay in proportions close to those observed in 1047 locally accrued cases (which serves as a model for the expected proportions of groups in a population based study) collected by the Haematological Malignancy Diagnostic Service (HMDS) in Leeds (Painter *et al*, 2018) and then by using a background derived from 52 samples generated with the HTG Pan B-Cell Lymphoma Panel assay in proportions close to those observed in 928 cases from the REMoDLB trial (Davies *et al*, 2019).

Table 8 shows the results. Agreement between the two was strong with 55 of 57 classifications identical. Figure 4 shows pairwise comparisons of the probability scores which are in high agreement (Pearson correlation GCB: r=0.9855, ABC: r=0.9852, UNC: r=0.9574).

**Table 8:** Comparison of classification of 57 DLBCLs from the Maple study using the standalone mode of DAC with two different background datasets; i) A set of 52 DLBCLs with proportions of ABC/GCB/UNC balanced according to proportions observed in the REMoDLB clinical trial ii) A set of 34 DLBCLs with proportions of ABC/GCB/UNC balanced according to locally accrued cases by the Haematological Malignancy Diagnostic Service (HMDS).

|  |  | Background, clinical trial |  |  |
| --- | --- | --- | --- | --- |
|  |  | ABC | GCB | UNC |
| Background data, population | ABC | 21 | 0 | 1 |
|  | GCB | 0 | 26 | 1 |
|  | UNC | 0 | 0 | 8 |

**Figure 4.**
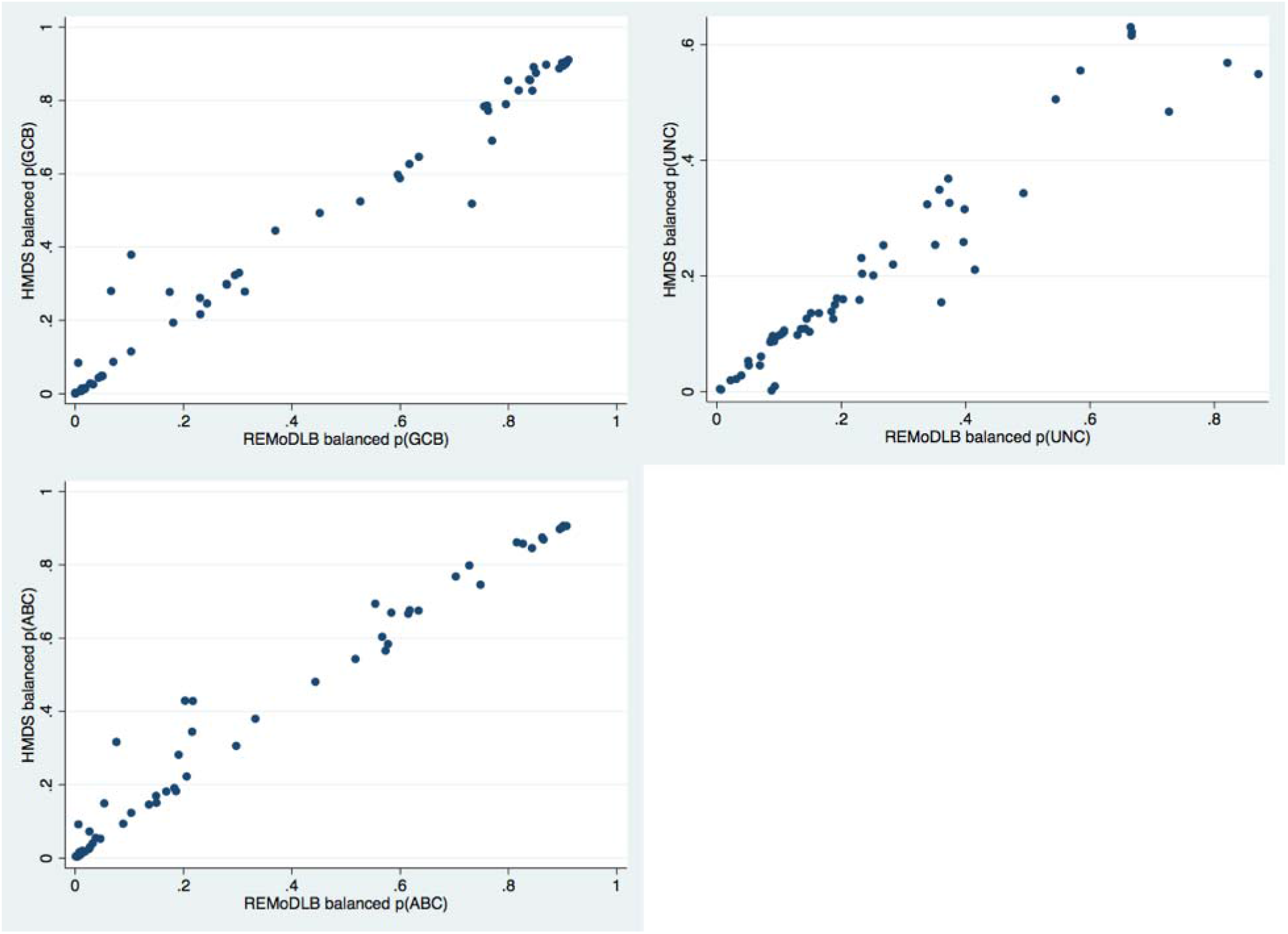
Comparison of probability scores for group membership to ABC/GCB/UNC for 57 DLBCLs from the MaPLe study. Agreement between the results is strong.

These results suggest that a background dataset with proportions reflecting a combination of those typically seen in clinical trial and population settings will work well for any new sample. Therefore, a finalised background dataset derived from 40 samples processed using the HTG Pan B-Cell Lymphoma Panel assay has been generated for prospective use.

### Using principal component analysis as an additional quality control step

For these data, quality control checks based upon checking negative/positive ratio, maximum raw read count, median read count and coefficient of variation were conducted, followed by principal component analysis as a further check to detect potential outliers. We wished to see how well the QC of the HTG EdgeSeq Reveal software agreed with this QC procedure. Thus, we conducted a comparative test of the QC results for 2 plates of 24 samples of the HTG Pan B-Cell Lymphoma Panel assay that contained 11 samples dropped during QC. Of these samples using the QC pipeline listed in the methods, 7 cases failed standard QC and 4 cases were removed as outliers based on principal component analysis. The HTG EdgeSeq Reveal software flagged 6 of the former cases but none of the PCA as failed samples. On further investigation, the cases that failed PCA represented specific subtypes of aggressive B cell lymphoma or had minimal tumour present. We conclude that principal component analysis may provide a useful additional method to identify potential outliers based on sample composition or disease subtype. Knowledge of the reason for being a PCA outlier is important, in order to decide if the sample(s) should be included in downstream analysis.

## Discussion

DAC is a highly flexible algorithm and has been shown to be able to carry out effective DLBCL Cell of Origin classification with gene expression profiles generated from a wide range of platforms (Ahmed *et al*, 2021). Here we have shown that the implementation of DAC in R gives highly comparable results compared to the original DAC implemented in Windows.

We have demonstrated that DAC-R COO classification of DLBCL samples processed using data from the HTG DLBCL COO panel provides comparable COO classification calls to those derived using the HTG DLBCL COO classifier. DAC-R can also be successfully applied to data derived from the HTG Pan B-Cell Lymphoma Panel and comparable results were obtained when compared to the original DAC classifications on Illumina WG-DASL data as well as using the HTG EdgeSeq Reveal DLBCL algorithm for probability of being GCB-type. Overall, concordance of all comparisons was at least 80%, which is similar to that reported in our previous studies (Ahmed *et al*, 2021). Most disagreements involved cases classified as UNC using DAC, a class that the HTG classifiers aims to minimise and which are preferentially classified as ABC or GCB by that algorithm. Other disagreements tended to have lower classification probability scores, indicating that they may represent borderline cases that lie close to classification boundaries, and probably reflecting biological heterogeneity and/or ongoing differentiation within individual tumours. Interestingly the survival difference of patients whose tumours are classified as ABC vs GCB is greater when only high classification probabilities are considered (Care *et al*, 2013). Therefore, consideration of the probability of classification is recommended where clinical application of COO classes is intended.

Whilst the DAC produced consistent COO results across different datasets, as well as in standalone and batch mode, the current study also highlights the requirement for the careful selection of background data which is used by DAC for data normalization. Changes to the background dataset can result in changes to classification, particularly in cases with lower probability of classification - so-called borderline or ‘edge’ cases. Using knowledge of a range of patient cohorts, we have derived a background dataset that is likely to be suitable for the COO classification of any DLBCL cohort.

COO classification remains the only gene expression signature used in the diagnosis of DLBCL in clinical practice, however additional gene expression profile subgroups including molecular high grade (MHG) (Sha *et al*, 2019) and DHITSig (Ennishi *et al, 2019*) have recently been defined. These groups are largely consistent between the two studies and add further subgroups beyond the COO, with both studies identifying a poor prognostic group within GCB.

The results of this study provide confidence in classification using gene expression profiling across different platforms, both in the classification of COO and, moving forward, with more recently described classification schema. The ability to use DAC with the HTG Pan B-Cell Lymphoma Panel enables users to make a COO classification as well as to use the extra genes on the panel to obtain additional information about the samples.

## Supporting information

TRIPOD checklist

## Data Availability

The REMoDLB data is presently available at GEO
The HMRN data is presently available at GEO
The MaPle data is part of an ongoing study and is not presently available at this time.

https://www.ncbi.nlm.nih.gov/geo/query/acc.cgi?acc=GSE117556

https://www.ncbi.nlm.nih.gov/geo/query/acc.cgi?acc=GSE181063

